# Heterogeneity in deaths of despair: excess mortality in the US during the Covid-19 pandemic

**DOI:** 10.1101/2024.01.14.24301291

**Authors:** Sasikiran Kandula, Katherine M. Keyes, Jeffrey Shaman

## Abstract

The impact of Covid-19 on mortality includes both direct effects of the virus and indirect effects mediated through other causal pathways. In the United States, the indirect effects, particularly from suicides, overdoses and alcohol-induced causes (i.e. deaths of despair) (1) are understudied. Here, we estimated excess non-Covid deaths and deaths of despair, in the US overall, in each state and in 72 demographic strata. Nationally, 114,230 (127,597) excess non-Covid deaths, 19,074 (33,559) excess poisoning deaths and 8,746 (13,771) excess alcohol-induced deaths were estimated during 2020 (2021). Excess poisoning and alcohol-induced mortality were highest among the 35-44 and the 55-64 year groups, respectively. The Black and the American Indian/Alaskan Native populations had the highest excess poisoning and alcohol-induced mortality, respectively. Fewer suicides than expected occurred nationally, but excess suicides were estimated among Black youth. These findings suggest that additional resources need to be mobilized to limit increases in deaths of despair.

## Introduction

The Covid-19 pandemic resulted in societal disruptions throughout the world. Assessment of the public health impacts of the pandemic must recognize both the direct effects of Covid-19 (morbidity and mortality following infection including long-Covid (2)), and its indirect effects including increased drug overdoses and impact on population mental health due to isolation and stress. Measures of morbidity from Covid-19 are sensitive to the healthcare system’s capacity to test and care for infected individuals. In the United States, as in many other countries, the healthcare system was reportedly overwhelmed during Covid-19 surges, introducing inaccuracies in case and hospitalization surveillance data. On the other hand, mortality reporting systems are more resilient to measurement and surveillance variance and are arguably a more reliable gauge of the *direct*, albeit more severe, effects of the pandemic. The mechanisms and pathways through which the pandemic has indirectly impacted population health are complex, but as is the case with direct effects, mortality is one of the more reliable measures of indirect effects of the pandemic.

Excess mortality, the difference between observed deaths from all causes during the pandemic and deaths expected in a counterfactual scenario in which a pandemic did not occur and mortality trends before the pandemic continued, encompasses both the direct and indirect effects of the pandemic and is a more comprehensive measure of its impact on public health. As an extension, the difference between expected and observed deaths from all causes other than Covid-19 can be interpreted as a measure of the indirect effect of the pandemic on mortality. Admittedly, deaths certified to be from causes other than Covid-19 can potentially include deaths from Covid-19 infections that were recorded as due to a different underlying cause in the absence of a confirmatory laboratory test for SARS-CoV-2 (3, 4). The prevalence of such mislabeled deaths and their variability over the course of the pandemic, across states and demographic groups is not quite known, although 91% of Covid-associated deaths (includes deaths for which Covid-19 infection was the presumed or probable contributing cause), were certified with Covid-19 as the underlying cause of death (5). With this caveat, excess deaths from non-Covid causes can be interpreted as a measure of indirect effects of the pandemic.

Multiple estimates of all-cause excess mortality in the US have been previously reported (6–15). These include estimates for the US overall, estimates at different geographical resolutions such as census regions or states, as well as for different racial and ethnic groups. In a few cases, excess deaths from specific causes and disease categories, as well as non-Covid causes, have also been reported (6, 11, 16).

In this study, using an extensive well-resolved historical record of mortality in the US, we estimated excess deaths from non-Covid causes to complement previously published estimates. A more important contribution, and one previously unreported to our knowledge, is a quantification of the pandemic’s impact on deaths of despair. Deaths of despair is a convenience label introduced by Case and Deaton and includes deaths from suicide, drug overdose and alcohol use (1, 17, 18). Case and Deaton have proposed that an underlying set of social and economic changes in the US — loss of meaningful work, shift in economic and political power from labor to capital, relaxation of social norms related to marriage, a diminished role of religion, inefficiencies in the healthcare system — are common factors impacting the rise in mortality from these causes, especially among middle-aged White men without a college degree.

Between 1999 and 2019, the age-adjusted mortality rate from poisoning in the US increased from 6.1 per 100,000 population to 21.6 (19, 20). Over the same period, suicides increased from 10.5 to 13.9, and alcohol-induced deaths among those 25 years or older increased from 10.9 to 16.0 (19, 21, 22). These increasing trends are anomalous as all-cause mortality during the same period has decreased from 875.6 to 715.2, as did mortality from some of the major leading causes of deaths such as heart disease (266.5 to 161.5), cancer (200.8 to 146.2), and stroke (61.6 to 37) (19).

With the emergence of the Covid-19 pandemic, there was widespread concern that the economic and psychological impacts of the pandemic would exacerbate these distal underlying drivers of deaths of despair. Large scale surveys conducted during the initial months of the pandemic indicated a general deterioration of population mental health with a significant proportion of those surveyed reporting depression, anxiety and thoughts of suicide (23). Similarly, the pandemic was expected to severely impact those with substance use disorders, either through disruptions in access to treatment or harm-reduction services, loss of contact with trusted drug sources of consistent potency, and relapses from pandemic related economic stress (24). Increased alcohol consumption as a coping mechanism was also hypothesized (25). Given the evidence of a concentration of deaths of despair among the middle-aged White non-Hispanic population before the pandemic, we examined whether excess mortality from deaths of despair during the pandemic had disproportionate representation from specific demographic groups.

With the help of a detailed dataset of all recorded deaths in the US between 2003 and 2019, we generated estimates of expected all-cause deaths, as well as deaths expected from poisoning, alcohol-induced causes and suicide in each county in the US during 2020 and 2021 (see *Methods*). Additionally, in order to capture group-specific temporal trends and spatial patterns leading up to the pandemic, we built separate models for 72 population strata (9 10-year age groups, 4 racial and 2 sex groups). These together enabled the generation of county- and strata-specific expected death estimates in each year.

By comparing the expected deaths with observed deaths, we were able to estimate excess mortality during the pandemic. Specifically, by comparing all-cause expected deaths with all deaths from causes other than Covid-19, we estimated the overall indirect effect of the pandemic on mortality. Similarly, by comparing expected deaths from each of suicide, poisoning and alcohol-induced causes with their respective observed deaths, we estimated the effect of the pandemic on each of these causes. While these estimates at the national level are informative, to further understand the differential impact of the pandemic by location and demographic group, we analyzed the variability in excess deaths across states, and by population age, race and sex..

## Results

### National excess mortality estimates from causes other than Covid-19

We have previously reported median all-cause excess deaths of 463,187 (95% uncertainty interval (UI): 426,139 – 497,526) in 2020 and 544,105 (95% UI: 492,202 – 592,959) in 2021 [in review, provided as supplementary text]. Excess deaths from causes other than Covid-19 were estimated to be 114,230 (95% UI: 57,999– 167,080) and 127,597 (95% UI: 75,709 – 176,447) during 2020 and 2021 respectively, with the corresponding excess mortality rates per 100,000 population at 36.8 (95% UI: 18.7 - 53.9) and 41.1 (95% UI: 24.4 – 56. 9).

Excess mortality rates from non-Covid causes among 35-44 year, 45-54 year and 55-64 year old populations were comparable to each other and to the national rate, and were higher among the 65-74 year age group (58.7 per 100,000 population) (Figure 1). Higher rates were observed in 2021 than in 2020 for all four of these age groups, with the 65-74 year group showing the largest increase and a near doubling of excess mortality (129.2; 95% UI: 69-189). Excess mortality rates among those under 15 years of age and over 75 years were not statistically significant, i.e. after excluding deaths certified to be from Covid-19, the observed deaths from all other causes were not statistically different from expected deaths from all other causes.

**Figure 1.**
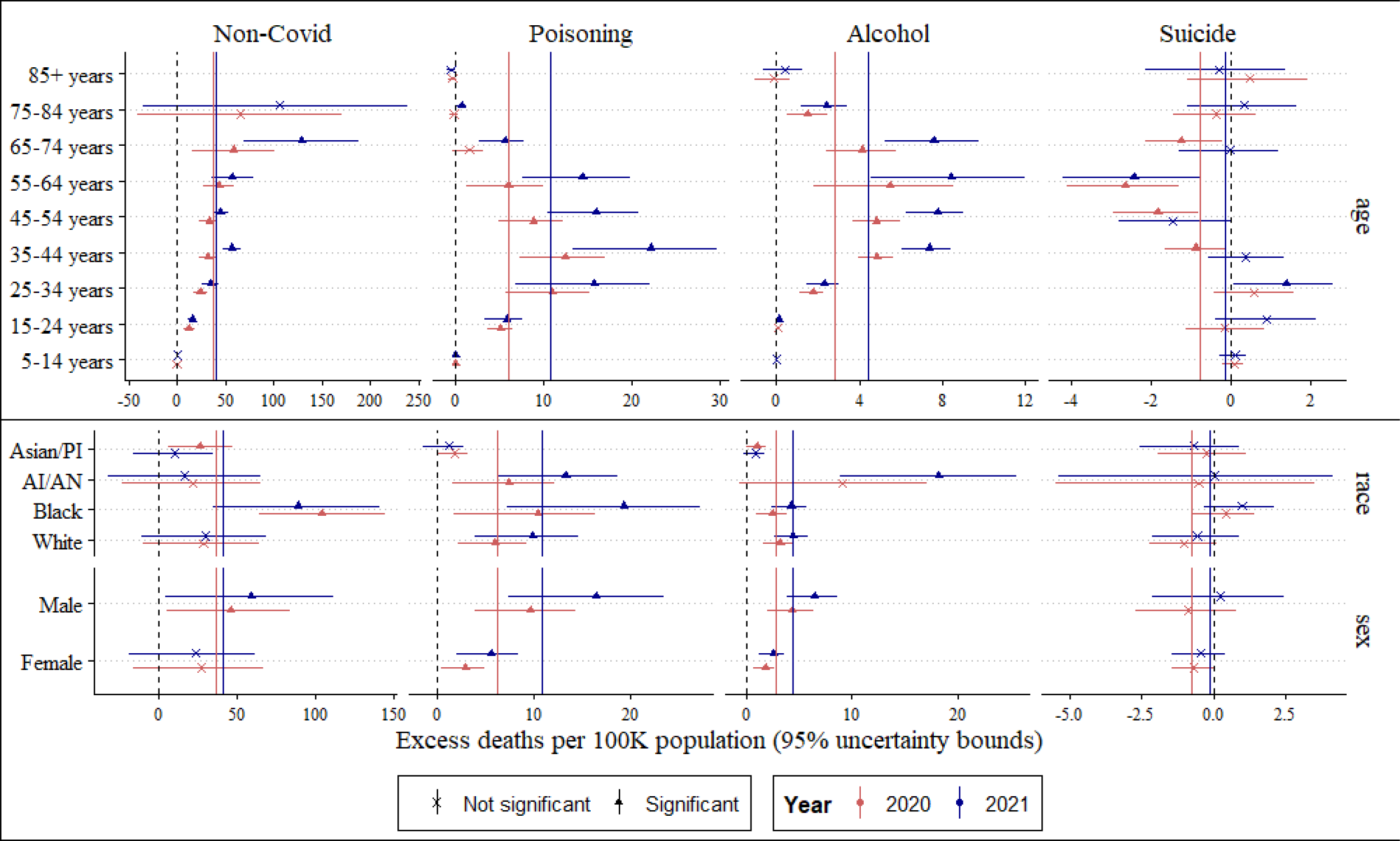
National excess mortality estimates in 2020 and 2021, stratified by age (top), race and sex (bottom). Estimates for race and sex are age-standardized to national age distribution using the direct method. The solid vertical lines denote national estimates. Instances where the lower and upper 95% uncertainty bounds had different signs were not considered to be significant (denoted by a *x*; for example *Non-Covid* in *5-14 years*). *Non-Covid* deaths in 85+ year group were not statistically significant and had a large uncertainty interval and hence suppressed to improve plot legibility.

Age-standardized excess mortality rates among women were also not statistically significant in either year. In contrast, among men, 46.3 excess non-Covid deaths per 100,000 population during 2020 and 59.5 during 2021 were estimated to have occurred. Further disaggregating by sex and age together showed statistically significant excess deaths among both men and women between 25 and 64 years of age, and higher excess mortality rates among men than in women in these age groups and during both years. The age distribution of excess deaths among men and women was similar in each year (Figure 2).

**Figure 2.**
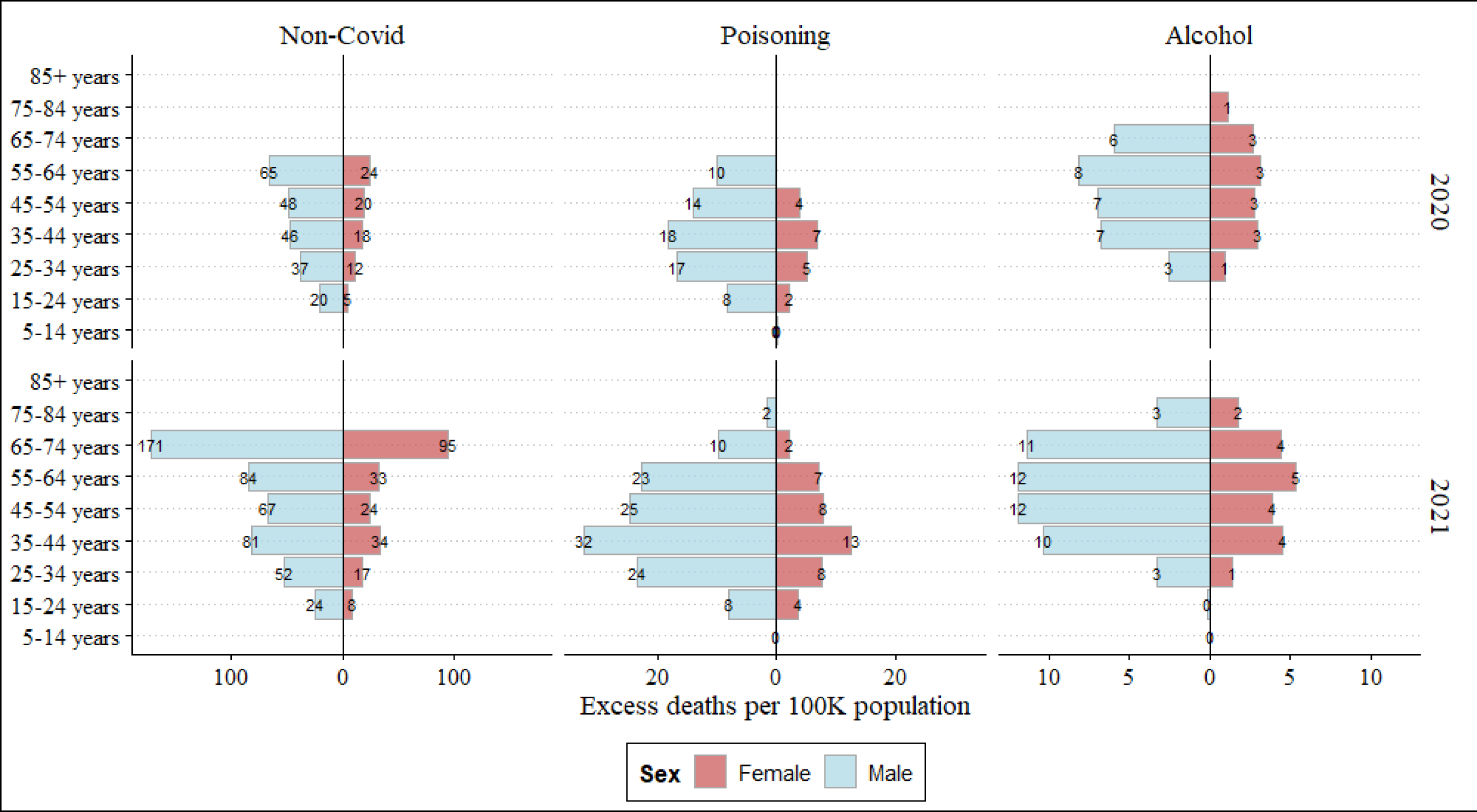
Excess mortality estimates from causes other than Covid-19 (Non-Covid), poisoning and alcohol-related causes, stratified by age and sex, during 2020 (top) and 2021 (bottom). Estimates not found to be statistically significant are suppressed. See Figure S1 for excess deaths from suicide.

Similarly, when stratified by racial groups, statistically significant excess deaths were estimated among the Black population (104. 5 during 2020 and 89.2 during 2021) and among the Asian/PI population during 2020 (26.8). Further disaggregating by race and age together showed excess deaths in the White, Black and AI/AN populations between 15 and 64 years (Figure 3). Among the 25-34 year and 35-44 year age groups, the AI/AN population had the largest non-Covid excess deaths of the four race groups.

**Figure 3.**
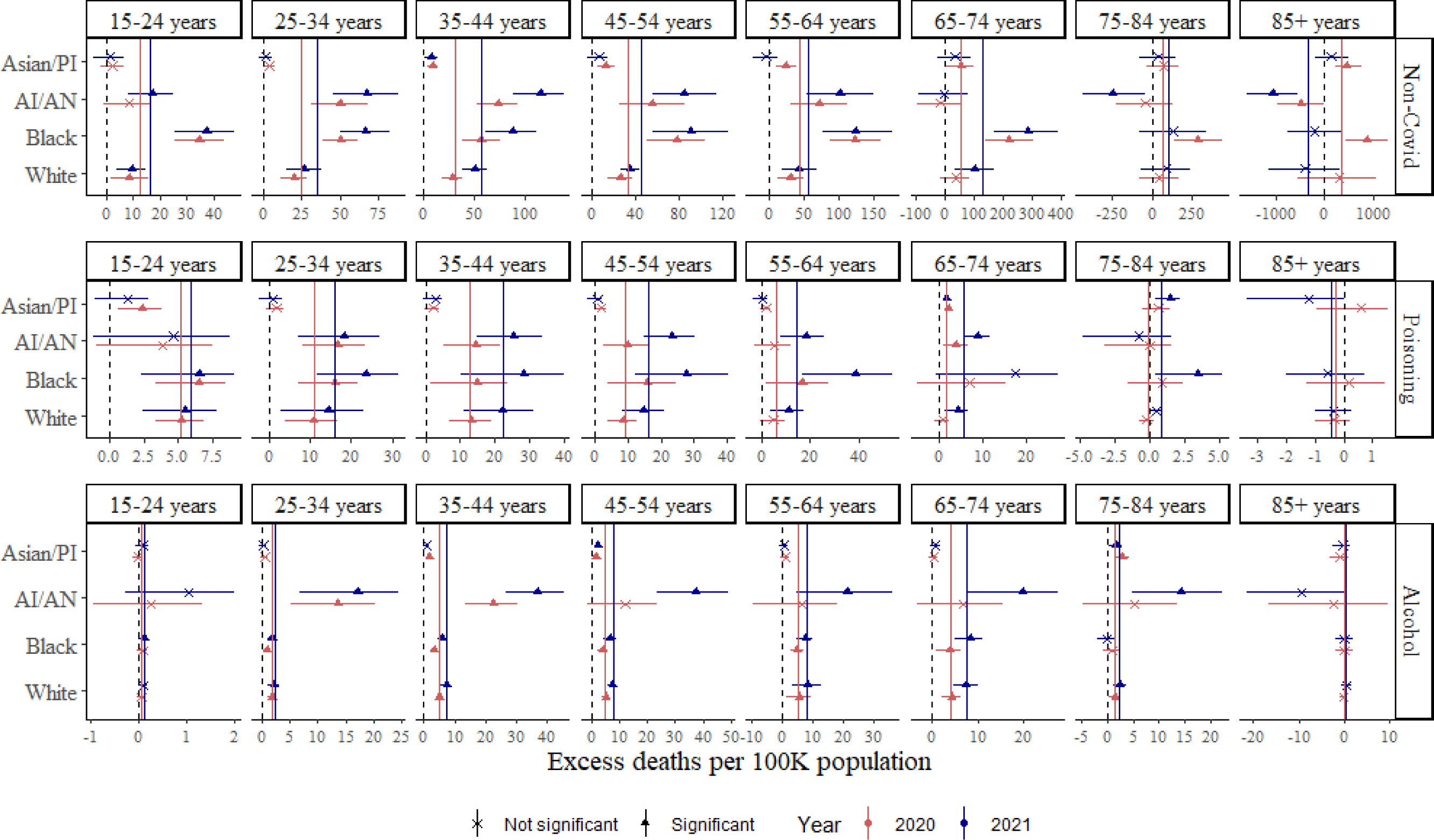
Excess mortality estimates from causes other than Covid-19 (Non-Covid), poisoning and alcohol-related causes, stratified by age and race, during 2020 (red) and 2021 (blue). Solid vertical lines denote excess mortality rate across all races. Instances where estimates were not statistically significant are denoted by *x*.

### National excess mortality estimates from poisoning

There were a reported 91,648 (106,484) deaths from poisoning in 2020 (2021)(26). Of these, 19,074 (95% UI: 14,980 – 23,200) during 2020 and 33,559 (95% UI: 27,232 – 39,607) during 2021 were estimated to be excess deaths, at a rate of 6.2 (95% UI: 4.8 – 7.5) per 100,000 population and 10.5 (95% UI: 8. 8 – 12.8), respectively (Figure 1). Note that these deaths were in excess of those expected from a continuation of the upward trend of poisoning deaths before the pandemic.

The age profile during both years was similar, with larger excess deaths estimated in the four 10-year age groups between 25-64 years. In both years, the 35-44 year age group had the highest mortality rate at nearly twice the national rate (12.5 in 2020 and 22.3 in 2021). Rates increased between 2020 and 2021 in all four of these age groups, with the largest percentage increase among the 55-64 year old group (6.1 in 2020 and 14.5 in 2021).

Age-standardized excess mortality rates from poisoning among men were nearly three times that of women in both years (2.9 among women and 9.6 among men in 2020; 5.6 and 16.4 respectively in 2021). Among both men and women, the 35-44 year population was estimated to have the highest excess mortality (7 among women and 18.2 among men in 2020; 12.6 and 32.3 respectively in 2021), with a greater separation from the remaining age groups in 2021 than in 2020 (Figure 2).

Age-standardized excess mortality rates stratified by race group showed the Black population to have the highest rates among the four race groups during both years (10.5 in 2020 and 19.2 in 2021), followed by the AI/AN population (7.4 and 13.3); in both cases rates nearly doubled between 2020 and 2021. Higher excess rates in the Black and AI/AN populations relative to the White population were consistent in all groups under 65 years, and were particularly elevated during 2021 in the 55-64 year (17.7 among White, 25.9 among AI/AN and 54.2 among Black populations) and 45-54 year age groups (20.9 among White, 30.5 among AI/AN and 40.4 among Black populations) (Figure 3).

### National excess mortality estimates from alcohol-induced causes

There were a reported 49,057 (54,255) alcohol-induced deaths in 2020 (2021) (26). Of these, 8,746 (95% UI: 6,978-10,266) deaths during 2020 and 13,771 (95% UI: 11,724 – 15,632) deaths during 2021 were estimated to be excess deaths, at a rate of 2.8 (95% UI: 2.3 – 3.3) per 100,000 in 2020 and 4.4 (95% UI: 3.8 – 5.0) in 2021 (Figure 1). As with excess mortality from poisoning, a similar age profile of excess deaths from alcohol-induced causes was estimated during both 2020 and 2021; however, the distribution skewed older than in poisoning, with a majority of excess deaths estimated to be in the four 10-year age groups between 35 and 74 years.

The difference between men and women was also smaller than was estimated for poisoning, but men still had more than twice as many excess deaths as women (1.8 in 2020 and 2.6 in 2021 among women; 4.4 in 2020 and 6.5 in 2021 among men). This difference was also consistent when disaggregated by age group and sex during both 2020 and 2021 (Figure 2).

Unlike poisoning, excess mortality among the Black population was comparable to that of the White population in both years, but excess mortality among the AI/AN population was particularly pronounced at nearly three times the national average in 2020, and four times the national average in 2021 (9.1 in 2020 and 18.2 in 2021). Furthermore, while in 2020 excess deaths among AI/AN were largely limited to the 25-44 year old population, in 2021 these extended to the older 45-84 year groups (Figure 3).

### National excess mortality estimates from suicides

In contrast to poisoning and alcohol-induced deaths, deaths from suicide were estimated to have decreased during the pandemic nationally. In 2020, 2,450 (95% UI: 1,427 to 3,567) *fewer* deaths from suicide were observed than were expected from pre-pandemic trends, at a rate of −0.79 (95% UI: −1.15 to −0.46) per 100,000 population. During 2021, the observed national suicide rate was not statistically different from expected rates. For reference, there were a reported 45,977 (48,178) suicide deaths in 2020 (2021) (26).

Disaggregating by age showed a statistically significant decrease in suicide rates during 2020 among 35-74 year age groups, with the largest decrease in the 55-64 year age group (−2.7 per 100,000). Suicide rates were higher in 2021 than in 2020 nationally, but a statistically significant difference was estimated for only two age groups: a deficit of 2.4 per 100,000 among the 55-64 year group, and an excess of 1.4 per 100,000 among the 25-34 year group (Figure 1).

Age-standardized excess rates for suicides following disaggregation by sex or race were not statistically significant. Disaggregating by age and sex together, however, showed significant deficits among middle-aged women (35-64 years) and 55-64 year men (Figure S1). Statistically significant excess mortality was also estimated when disaggregated by age and race together, with deficits in the above age groups largely limited to the White population, and *excess* suicides estimated among the younger Black population (1.8 in 2020 and 2.9 in 2021 among 15-24 year group; 2.5 in 2021 among 25-34 year group).

### Excess mortality estimates for states

We further estimated excess mortality rates from all causes other than Covid-19, as well as deaths of despair, in the 50 states and the District of Columbia. Statistically significant excess mortality from non-Covid causes were estimated in 37 states during 2020, and in 39 states during 2021. Of the 31 states with statistically significant estimates in both years, an increase between 2020 and 2021 was estimated in 21 states.

The same five states — Mississippi (108.3 excess deaths per 100,000 population in 2020; 116.8 in 2021), West Virginia (106; 180.4), Kentucky (83.1; 108.2), New Mexico (81.6; 122.3) and Tennessee (80.5; 109) — were estimated to have the five highest non-Covid excess deaths in each of 2020 and 2021; in all 5 states, the excess mortality rate was higher in 2021 than in 2020 (Figures 4, 5). The Appalachian states of West Virginia, Kentucky and Tennessee were also estimated to have high excess mortality from poisoning, while New Mexico had large excess mortality from alcohol-induced causes. In Mississippi, excess mortality from both poisoning and alcohol-induced causes were not high relative to other states, suggesting contributions from causes other than those that were the focus here.

**Figure 4.**
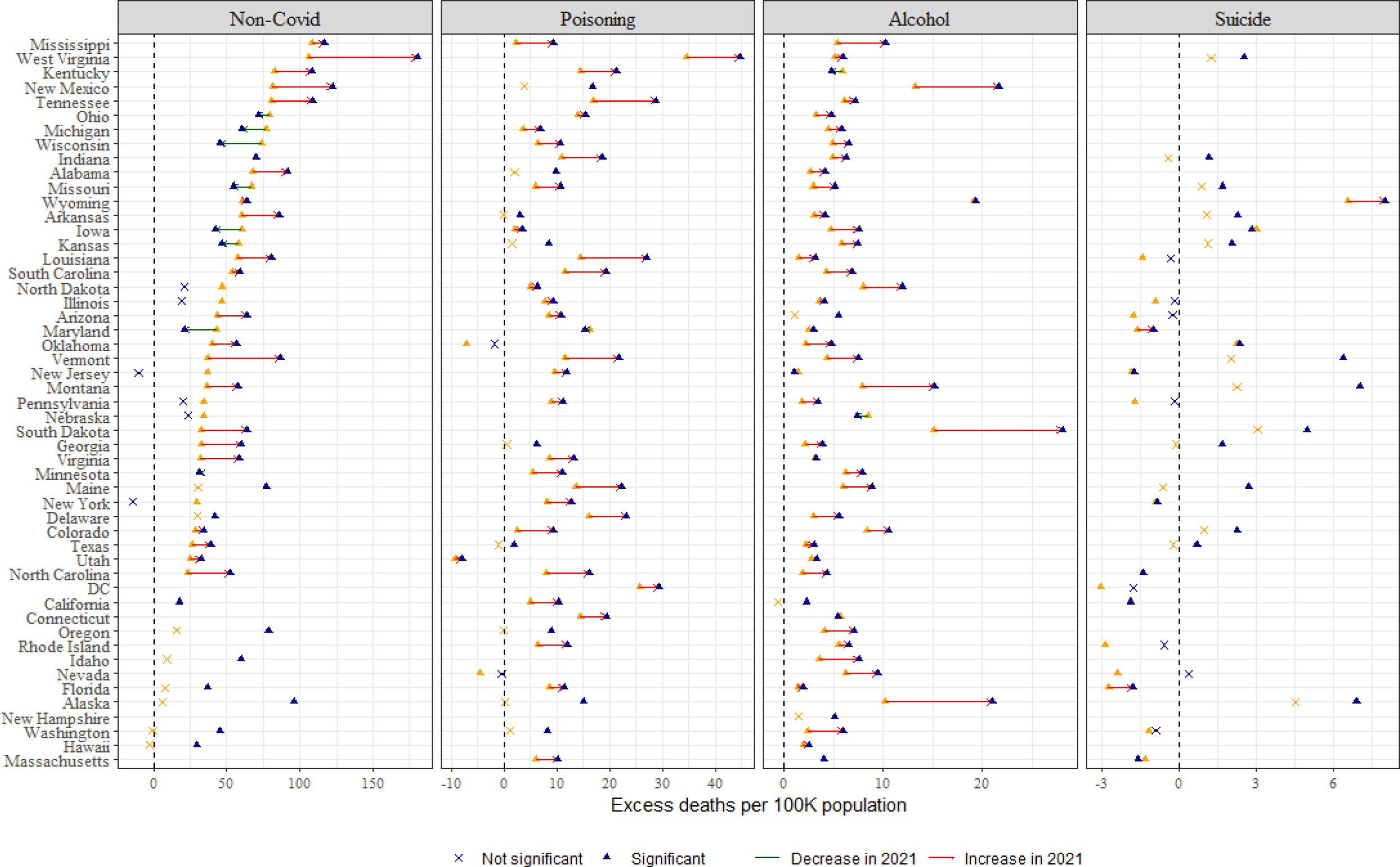
Change in state-level excess mortality rate from causes other than Covid-19 (Non-Covid, left), poisoning, alcohol-related causes and suicide between 2020 (orange data point) and 2021 (blue data point). Estimates that were not statistically significant are denoted by *x*, and states that were not significant in both years were suppressed (for example, Suicides in Mississippi). Change in excess rate between 2020 and 2021 is indicated by the color of the arrow.

**Figure 5.**
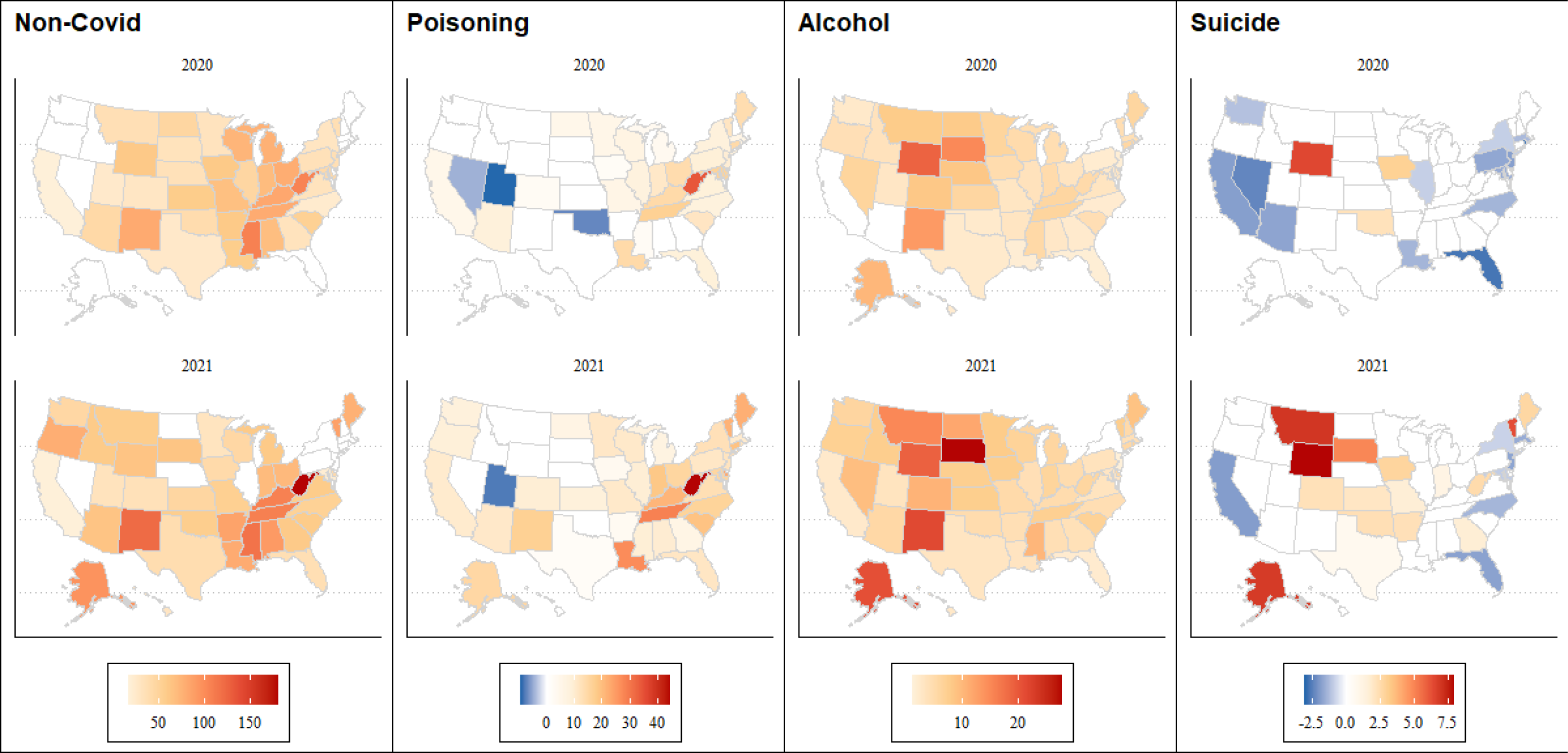
State-level excess mortality rate from causes other than Covid-19 (Non-Covid), poisoning, alcohol-related causes and suicide, during 2020 (top) and 2021 (bottom). Estimates that were not statistically significant were assigned no color. The magnitude of excess (deficit) rates is represented by varying shades of red (blue).

Statistically significant excess mortality from poisoning was observed in two-thirds of the states during both years, with nearly all of these states indicating an increase in 2021 from 2020. In addition to the Appalachian cluster noted above, the District of Columbia (25.7 in 2020; 29.3 in 2021), Louisiana (14.4; 27.1) and four states in the northeast — Delaware (16.1; 23.2), Maine (13.7; 22.2), Connecticut (14.4; 19.5) and Vermont (11.6; 21.9) — were estimated to have experienced large excess deaths from poisoning during both years.

While the magnitude of excess mortality from alcohol-induced causes was smaller than from poisoning, these were more widespread with excess mortality indicated in 46 states during both years. In addition to New Mexico noted above, a cluster of states in the Mountain region — South Dakota (15.1 in 2020; 28.1 in 2021), Wyoming (19.1; 19.3), Montana (7.9; 15.2) and North Dakota (8; 11.9) — along with Alaska (10.2; 21) had high excess mortality rates from alcohol-induced causes. Particularly large increases between 2020 and 2021 were noted in South Dakota and Alaska (Figure 4).

As previously noted, fewer suicide deaths than expected occurred during 2020 in the US nationally. Disaggregating by state showed statistically significant deficits in 15 states, with the largest deficits in the District of Columbia (−3.1), Rhode Island (−2.9), Florida (−2.8) and Nevada (−2.4). Observed suicide rates were not statistically different from expected rates in 33 states.

On the other hand, while nationally expected suicides were in line with observed suicides during 2021, at the state-level excess deaths were estimated for 16 states. Wyoming was a clear outlier with the highest excess suicide deaths in both years (6.6 in 2020; 8.1 in 2021). Montana (7.1), Alaska (6.9), Vermont (6.4) and South Dakota (5), which were identified as having had high excess deaths from alcohol-induced causes, were also estimated to have high excess suicides during 2021. Besides Wyoming, Iowa (3; 2.8) and Oklahoma (2.2; 2.4) were the only states estimated to have excess suicides in both years. Six states with sizeable urban populations — California, Maryland, Massachusetts, New York, New Jersey and North Carolina — had fewer than expected suicides in both years.

## Discussion

Our estimates show that mortality burden was an additional 32.6% and 30.6% above deaths reported with Covid-19 as underlying cause, during 2020 and 2021 respectively. While a majority of Covid-19 deaths have occurred in the 75+ population (58.8% in 2020 and 43.1% in 2021), indirect deaths were largely limited to those under 75 years. The Black and AI/AN populations were reported to have greater mortality from Covid-19 than the White population, and the indirect effects in these two groups were also estimated to be larger than in the White population. Geographical variability of Covid-19 deaths and non-Covid excess deaths was also observed (not reported); these findings together indicate that assessments of the pandemic’s overall mortality burden would be incomplete without the inclusion of excess non-Covid deaths.

Our results also show that nationally excess deaths from suicide, poisoning and alcohol-induced deaths together accounted for 25% (36%) of excess non-Covid deaths during 2020 (2021), indicating a sizeable contribution from causes other than deaths of despair. Limited access to emergency services following a heart attack (27) or stroke (28), changes in health-seeking behavior to avoid infection in clinical settings (29, 30), changes in patient care protocols to prioritize Covid-19 cases (leading to fewer referrals for suspected cancer (31), for example) could have contributed to excess mortality. Of note, excess non-Covid mortality was particularly elevated among the 65-74 year population, who are more likely to have chronic conditions requiring regular access to care than younger age groups and for whom the above factors could have played a greater role. Additionally, excess non-Covid mortality was larger among the Black population than in all other racial groups, and the extent to which this is due to differences in the prevalence of chronic conditions and the uneven indirect socioeconomic impact of the pandemic among racial groups remains to be investigated.

Poisoning deaths remained relatively stable between 2017 and 2019 at nearly 70,000 deaths per year. This followed a 10% year-to-year increase between 2014 and 2016 primarily due to increased availability of fentanyl and other synthetic opioids. Usage patterns of illicit drugs have changed periodically over the last two decades with a transition from prescription opioids to heroin to synthetic opioids, and more recently towards (meth)amphetamines (20), with consequent effects on overdose deaths. It is difficult to disentangle the extent to which the excess poisoning deaths in 2020 and 2021 were due to another shift in usage driven by supply-side dynamics which would have occurred even without a pandemic, or an indirect effect of the pandemic, or more likely, a compounded effect of the two.

However, while poisoning deaths before the pandemic were predominantly among the middle-aged White population, excess poisoning deaths during the pandemic were estimated to be at a higher rate in younger age groups (25-45 years), and higher among the Black population than the White population, indicating a possible shift in overdoses to a younger non-White population during the first two years of the pandemic.

Most states implemented policies to mitigate anticipated spikes in overdoses during the pandemic, and the excess mortality estimates reported here can help evaluate the efficacy of these state-specific measures, especially when large differences are observed between adjacent states. For example, West Virginia, the District of Columbia and Louisiana had higher excess mortality from poisoning relative to their neighbors and are suitable candidates for a policy comparison. Efforts to reduce overdose through increased naloxone availability, reducing high-risk opioid prescribing, and supporting and building treatment capacity for substance use disorders are all evidenced-based approaches for addressing the continued drug poisoning epidemic. Such efforts may need to be enhanced and further supported through funding and policy change especially in these states.

Unlike excess poisoning mortality, the age distribution of excess alcohol-induced mortality during the pandemic was similar to the age distribution of alcohol-induced mortality during the pandemic, and no shift in burden towards younger populations was detected. However, there was a possible proportional shift in alcohol-induced mortality from the White population to both AI/AN and Black populations during 2021. Excessive alcohol use was estimated to have accounted for 1 in 8 deaths in the US population aged 20 to 64 during the 5-year period leading up to the pandemic (32) and further increased by nearly 25% during the first year of the Covid-19 pandemic (33). Together with excess estimates reported here, these results suggest substantial burden of alcohol-induced deaths in middle-age and older populations and need for alcohol regulations to reduce mortality.

In light of mental health surveys conducted during the initial months of the pandemic on suicidal thoughts, plans or attempts which indicated a possible uptick in suicides, the decrease in observed suicide rates during 2020 was a positive public health outcome. However, caution is required in interpreting the deficit suicide estimates reported here. Out-of-sample validation of our model’s expected mortality estimates during pre-pandemic years had a positive bias (see *Methods*). Although this would lead to a more conservative excess death estimates for poisoning and alcohol-induced causes, due to the decreasing trend in suicide from 2018, this bias could have contributed to an erroneous estimation of a deficit in suicides during 2020. On the other hand, and for the same reason, excess suicides in the 15-24 year Black population in 2020 and the 15-34 year Black population in 2021 are likely to be underestimates and additional analysis is necessary to understand the underlying causes for the increase in suicides among Black youth (Figure S1).

The causal pathways for fewer suicides than expected during the pandemic in states with large urban centers is unclear, but a combination of the ‘pulling together’ effect (34) and the alleviation of the impact of a strained social safety net through pandemic stimulus checks, eviction moratoriums and related measures may be at play.

Limitations of this study include the use of provisional estimates of mortality for 2021 which could have a larger effect on minority groups (see Appendix Text 1); an inability to age-standardize excess mortality rates at the state-level; inaccuracies in cause of death certification both for Covid-19 and poisoning; concerns about intentionality certification of overdose deaths (35, 36); use of 2020 population estimates for 2021 without adjusting for the high mortality in 2020 (particularly impacting the elderly); and, large uncertainty bounds in model estimates for strata with small and sparse population.

Overall, we estimate that over 70,000 excess deaths of despair occurred during the first two years of the Covid-19 pandemic in the United States. Poisonings and alcohol-induced deaths as well as excess mortality from non-Covid causes exhibited considerable heterogeneity by demographic characteristics and across states. These findings highlight the indirect health burden of the pandemic, the need for further study of these outcomes and more targeted interventions to limit mortality.

## Materials and Methods

### Mortality

All-cause deaths for years 2003-2020 were obtained from a detailed mortality dataset from the US National Vital Statistics System, along with decedent age at time of death, race, sex, and county of residence (19). From these records, county- and strata-specific annual death counts were computed, where a strata was defined by a combination of decedent age, race and sex. 9 age groups (5-14 years, 15-24, …, 75-84 years, 85+ years), 4 racial groups (White, Black, American Indian/Alaskan Native (AI/AN) and Asian/Pacific Islander (Asian/PI)) and two sex groups (Male and Female) were used for a total of 72 strata. Annual county-level mortality rates (deaths per 100,000 population), were calculated with annual strata-specific county population estimates from the Bridged-Race Intercensal (2003–2009), decennial census (2010) and Postcensal (2011–2020) datasets (37, 38).

Cause-specific mortality from Covid-19, suicides, poisoning and alcohol-induced deaths were identified using *International Classification of Diseases-10 (ICD-10)* (*39, 40*) underlying cause of death codes (Table S1), and aggregated by county and demographic strata.

As detailed mortality data were not available for 2021 at the time this analysis was undertaken, provisional death counts and rates (not stratified) were retrieved from the Multiple Cause of Death public interface on CDC WONDER (26). The interface suppressed counties with fewer than 10 deaths resulting in the suppression of 14% of counties for Covid-19, 68% of counties for suicides and alcohol-induced deaths, and 59% of counties for poisoning. Consequently, estimation of excess deaths (difference from observed) was not possible at the county-level in 2021, and we limit to reporting estimates for states and the US overall.

### Bayesian models of expected deaths

To estimate expected all-cause deaths, we trained 72 strata-specific models using annual county-level deaths for the population in each strata between 2003 and 2019. Analogously, expected cause-specific mortality was estimated by training the models with annual cause-specific deaths. For simplicity, we limit following description to all-cause deaths.

Let *y*_*tsk*_ denote the number of all-cause deaths during year *t* in county *s* among the population in the *k^th^* strata, defined by a combination of age, race and sex of the decedent; *P*_*tsk*_ and *r*_*tsk*_ the population and risk of *k^th^* strata respectively. Under the assumption of a Poisson distribution for *y*_*tsk*_, risk is modeled as:

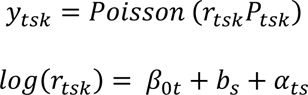

Here, *β*_0*t*_ is the time specific intercept calculated as *β*_0*t*_ = *β*_0_ + *ε*_*t*_, *β*_0_ representing the global intercept. *ε*_*t*_, the random effect representing the deviation of each year from *β*_0_, was modeled as a first-order random walk process with hyperparameter *τ*_*ε*_ (as described below):

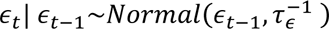

The Besag-York-Mollie (41) model was used to model the spatial random effect *b*_*s*_ as

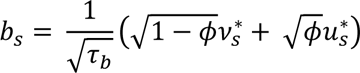

where 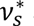 and 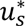 represent standardized (variance equal to 1) unstructured random effect (*v*_s_) and spatially structured effect (*u*_*s*_), respectively; 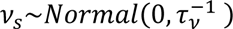 and

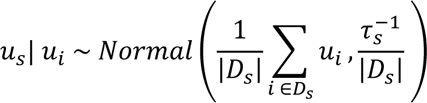

Here, *D*_*s*_ denotes the set of neighboring counties of county *s*, |*D*_*s*_| the number of neighboring counties, and *ϕ* a mixing parameter (0 ≤ *ϕ* ≤ 1) controlling the proportion of marginal variance explained by the structured effect. *α*_*ts*_ denotes a space-time interaction modeled using independent and identically distributed Normal prior as 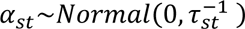.

Penalizing complexity (PC) priors were specified for hyperparameters *τ*_*ε*_, *τ*_*b*_, *τ*_*s*_, *ϕ* and *τ*_*st*_ (42). To impose an upper bound on spatial relative risk based only on structured or unstructured variation at *exp*(2), the prior for 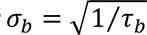 was specified as *Pr*(*σ*_*b*_ > 1) = 0.01 (43, 44). For mixing parameter *ϕ*, as the relative contribution of the unstructured or structured effects to *b* is uncertain, a broad prior was specified with *Pr*(ϕ < 0.5) = 0.5. Priors for all other hyperparameters are identical to that of *τ*_*b*_. Prior for the fixed effect *β*_0_ was specified with a minimally informative Normal distribution.

### Posterior estimates and aggregation

To get estimates of expected deaths for each of 2020 and 2021 from each of the *k* models, Monte Carlo simulations were generated and 1000 random samples were drawn from the joint posterior distribution:

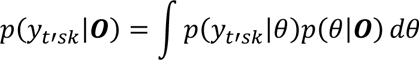

where *θ* is the (hyper) parameter vector (*β*_0_, *u*_*s*_, *v*_*s*_, *τ*_*ε*_, *τ*_*b*_, *τ*_*s*_, *ϕ* and *τ*_*st*_), and ***O*** the observed data through 2019.

The Integrated Nested Laplace Approximation (INLA) approach, as implemented in the *INLA* package (45, 46) in R (47), was used for Bayesian inference. To obtain posterior distributions of expected deaths for US nationally posterior distributions of all 72 models were aggregated. To obtain posterior distributions for a state, posterior distributions from all 72 models for the counties in the state were aggregated. To obtain estimates for a specific population subgroup (age, race or sex), relevant subset of the *k* models was identified, and posterior distributions from this subset were aggregated. Location- and strata-specific estimates were obtained analogously with a selection of relevant counties and models. For example, to estimate expected deaths among the White population in New York, the 18 models (9 age and 2 sex groups) in which race of the decedent was *White* were identified, and posterior distributions of expected deaths in only the counties in New York were summed.

Distributions of all-cause *excess* deaths were calculated as the difference between observed deaths and posterior distributions of expected all-cause deaths. Distributions of excess deaths from causes other than Covid-19 *(non-Covid)* were calculated by further subtracting Covid-19 deaths (deaths explicitly coded with U07.1 as the underlying cause). Mean and 95% uncertainty bounds were computed from the posterior distributions. An estimate was considered to be significant if the lower (2.5%) and upper (97.5%) bounds had the same sign.

Distributions of cause-specific excess deaths were calculated as the difference between observed deaths from the specific cause and posterior distributions of expected cause-specific deaths. To reiterate, the posterior distributions of each of the expected all-cause deaths, and expected suicides, poisoning and alcohol-induced deaths were from 4 distinct sets of 72 county-level strata-specific models, each set of models trained on annual observed deaths for the specific cause during 2003-2019.

To adjust for differences in age profiles of the different population sub-groups. the race and sex disaggregated excess mortality rates for the US nationally, as well as the state-level rates, were age-standardized using the direct method (48, 49), with national age distribution as the reference population (see Appendix Text 2).

### Temporal cross validation

As a temporal cross-validation exercise, for each of the years 2010-2019, we trained the models with data up to (and not including) the candidate year and projected deaths one year ahead. A mismatch between expected and observed deaths does not necessarily indicate sub-optimal quality of the models, but rather possible excess deaths from other factors. Inspecting the concurrence between the observed and predicted deaths could help detect consistent biases in model estimates.

Figures S3 a-d show uncertainty bounds of the expected mortality rates against observed rates for all cause deaths, poisoning, alcohol-induced deaths and suicides respectively, in the US overall, and disaggregated by age, race and sex. National expected death estimates were within the 95% uncertainty bounds in 7 of the 10 years. Cause-specific expected death estimates were biased higher, suggesting the excess death estimates for 2020 and 2021, are potentially underestimates. Similar patterns were observed in estimates of all-cause deaths for men and women and in 3 of the 4 race groups. Estimates for AI/AN were biased higher in a majority of the years, possibly due to the relatively sparse distribution of AI/AN population but this remains to be investigated. Cause-specific race and sex disaggregated estimates also at times showed positive bias.

See Appendix Text 3 for a comparison of excess non-Covid mortality estimates from the method described here and those estimated by the National Center for Health Statistics using a different method.

## Supporting information

Supplementary text and figures combined

## Data Availability

All data produced in the present study are available upon reasonable request to the authors

## Acknowledgements

This work is supported by a grant from the US National Institute of Mental Health (R01-MH121410), a contract from the US Centers for Disease Control and Prevention (75D30122C14289), and a gift from the Morris-Singer Foundation.

## Competing Interests

JS and Columbia University declare partial ownership of SK Analytics. JS consulted for BNI. KMK and SK have no competing interests.

## Data and code availability

Excess mortality estimates from this study for 2020 and 2021, and a data dictionary are included as supporting information.

