## Supplementary text and figures combined for "Heterogeneity in deaths of despair: excess mortality in the US during the Covid-19 pandemic"

*Supplementary information*

Sasikiran Kandula[1*], Katherine M. Keyes[2], Jeffrey Shaman[1,3]

[1]Department of Environmental Health Sciences, Columbia University, New York, NY.

[2]Department of Epidemiology, Columbia University, New York, NY.

[3]Columbia Climate School, Columbia University, New York, NY.

*Corresponding author:

*Appendix Text 1: Mismatch in race categories*

The race variable employed in the detailed mortality dataset for the years 2003 to 2020 used four categories: White, Black, American Indian/Alaskan Native (AI/AN) and Asian/Pacific Islander (Asian/PI). For 2021, for which the detailed dataset was unavailable at the time the analysis was undertaken, and provisional data from CDC Wonder Multiple cause of death (MCD) interface were used, the only available race variable used five categories with an additional category called 'more than one race'.

In other words, decedents who would be assigned to one of the four race categories in the detailed mortality dataset were assigned to the 'more than one race' category in the MCD. To assess the impact of this difference in categorization, we examined aggregate counts for the 4 race groups under the two variables during 2020 for all-cause deaths as well as 4 specific causes (see Table). We found that while under 1% of the White decedents were reclassified as 'more than one race' by the MCD, 8.1% of AI/AN decedents and 4.4% of Asian/PI decedents were classified as 'more than one race'. This high percentage of reclassification among AI/AN and Asian/PI was also seen in cause specific deaths, suggesting that interpretation of the change in excess rates between 2020 and 2021 among all race groups except White merits caution.

| **Race group** | **All-cause, Diff (%)** | **Covid-19, Diff (%)** | **Poisoning, Diff (%)** | **Alcohol, Diff (%)** | **Suicide, Diff (%)** |
|---|---|---|---|---|---|
| White | 7717 (0.28) | 566 (0.21) | 560 (0.77) | 217 (0.52) | 330 (0.82) |
| Black | 2665 (0.59) | 193 (0.33) | 343 (2.15) | 59 (1.32) | 147 (4.15) |
| AI/AN | 2281 (8.07) | 205 (4.37) | 144 (11.6) | 87 (4.5) | 83 (11.6) |
| Asian/PI | 4404 (4.36) | 307 (2.21) | 232 (17.5) | 75 (10.1) | 141 (8.99) |
| More than one | 17067 (NA) | 1271 (NA) | 1279 (NA) | 438 (NA) | 701 (NA) |

*Appendix text 2. Age standardization*

When comparing mortality rates across time or between groups, it is necessary to ensure that the age composition in the two groups is similar. In the US, there are known differences in age distributions across states (for example, age distribution skews older in Florida compared to most other states), by sex (women live longer than men) and race. As such, age standardization was required in this study to control for differences in age distributions and to render comparisons meaningful. The direct method of age standardization computes the mortality rates that would occur in a comparison group (say a state) if the observed age-specific death rates in the group were present in a population with the same age distribution as a reference population. Here, we used the US national age distribution in 2020 as the reference population.

The direct method has been implemented in the *epitools* package (1) in *R* (2) based on a method described by Anderson and Rosenberg (3). Briefly, using state excess mortality rates as an example, let $p_s^a$ denote the population in age group $a$ and state $s$ and $d_s^a$ the corresponding excess deaths. Age-specific excess mortality rate in state $s$ for age group $a$ is hence $d_s^a/p_s^a$, and the unadjusted mortality rate for state s is given by $\frac{\sum_a d_s^a}{\sum_a p_s^a}$. The state's age-standardized mortality rate is given by $\sum_a \left(\frac{d_s^a}{p_s^a} * \frac{\sum_s p_s^a}{\sum_{a,s} p_s^a}\right)$, which can be seen as a weighted average where the weight, $\frac{\sum_s p_s^a}{\sum_{a,s} p_s^a}$, is the proportion of the national population ($\sum_{a,s} p_s^a$) that belongs to age group $a$ ($\sum_s p_s^a$).

*Appendix text 3*: *Comparison with NCHS excess mortality estimates for causes other than Covid-19*

For all causes other than Covid-19 (i.e. Non-Covid), we compared national and state excess mortality estimates from the method described here with estimates published by the National Center for Health Statistics (NCHS) (4). National NCHS' estimates, at 16.8 and 26.7 excess deaths per 100,000 population in 2020 and 2021, respectively, were lower than our estimates (36.8 and 41.1). At the state level, between the two excess estimates a Spearman's correlation of 0.45 in 2020 and 0.79 in 2021 was estimated. NCHS' estimates were lower than our estimates in two-third of the states in both years (Figure S2).

In 2021, the largest differences between NCHS' and our estimates were in states where we predicted the largest excess deaths with overlap from poisonings and alcohol-related causes, namely West Virginia, Kentucky, Tennessee, New Mexico and Mississippi. In addition to these states, sizeable differences were noted in Ohio, Indiana and Wyoming during 2020.

| Underlying cause of deaths | ICD-10 codes |
|---|---|
| Covid-19 | U07.1 |
| Suicide | X60-X84, Y87.0, U03 |
| Poisonings | *X40-X44* (Accidental poisoning by and exposure to drugs, medicaments and biological substances), *X60–X64* (Intentional self-poisoning by and exposure to drugs, medicaments and biological substances), *X85* (Assault by drugs, medicaments and biological substances), and *Y10–Y14* (Poisoning by and exposure to drugs, medicaments and biological substances, undetermined intent). |
| Alcohol-induced deaths | *E24.4* (Alcohol-induced pseudo-Cushing's syndrome), *F10.** (Mental and behavioral disorders due to alcohol use), *G31.2* (Degeneration of nervous system due to alcohol), *G62.1* (Alcoholic polyneuropathy), *G72.1* (Alcoholic myopathy), *I42.6* (Alcoholic cardiomyopathy), *K29.2* (Alcoholic gastritis), *K70* ( Alcoholic liver disease), *K85.2* (Alcohol-induced acute pancreatitis), *K86.0* (Alcohol-induced chronic pancreatitis), *R78.0* (Finding of alcohol in blood), *X45* (Accidental poisoning by and exposure to alcohol), *X65* (Intentional self-poisoning by and exposure to alcohol), and *Y15* (Poisoning by and exposure to alcohol, undetermined intent) |

**Table S1**. ICD-10 underlying cause of death codes used to identify deaths from Covid-19, suicides, poisonings, and alcohol-induced deaths (5).

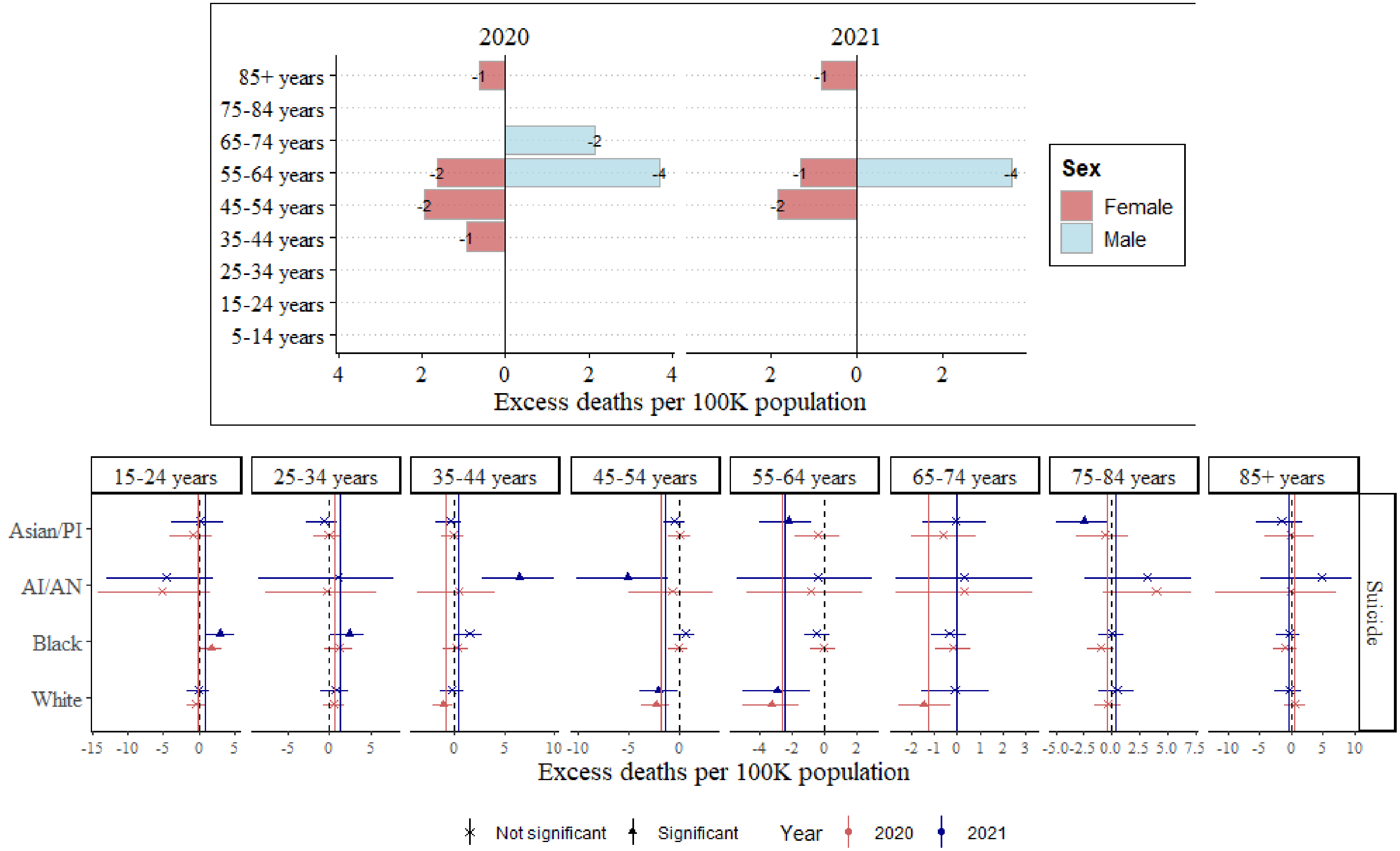


**Figure S1.** Excess mortality estimates from suicides: (*top*) stratified by age and sex showing statistically significant estimates only. All estimates are deficits (i.e. observed deaths lower than expected); (*bottom*) stratified by age and race. Estimates not statistically significant are denoted by *x* and solid vertical lines show excess estimate across all races

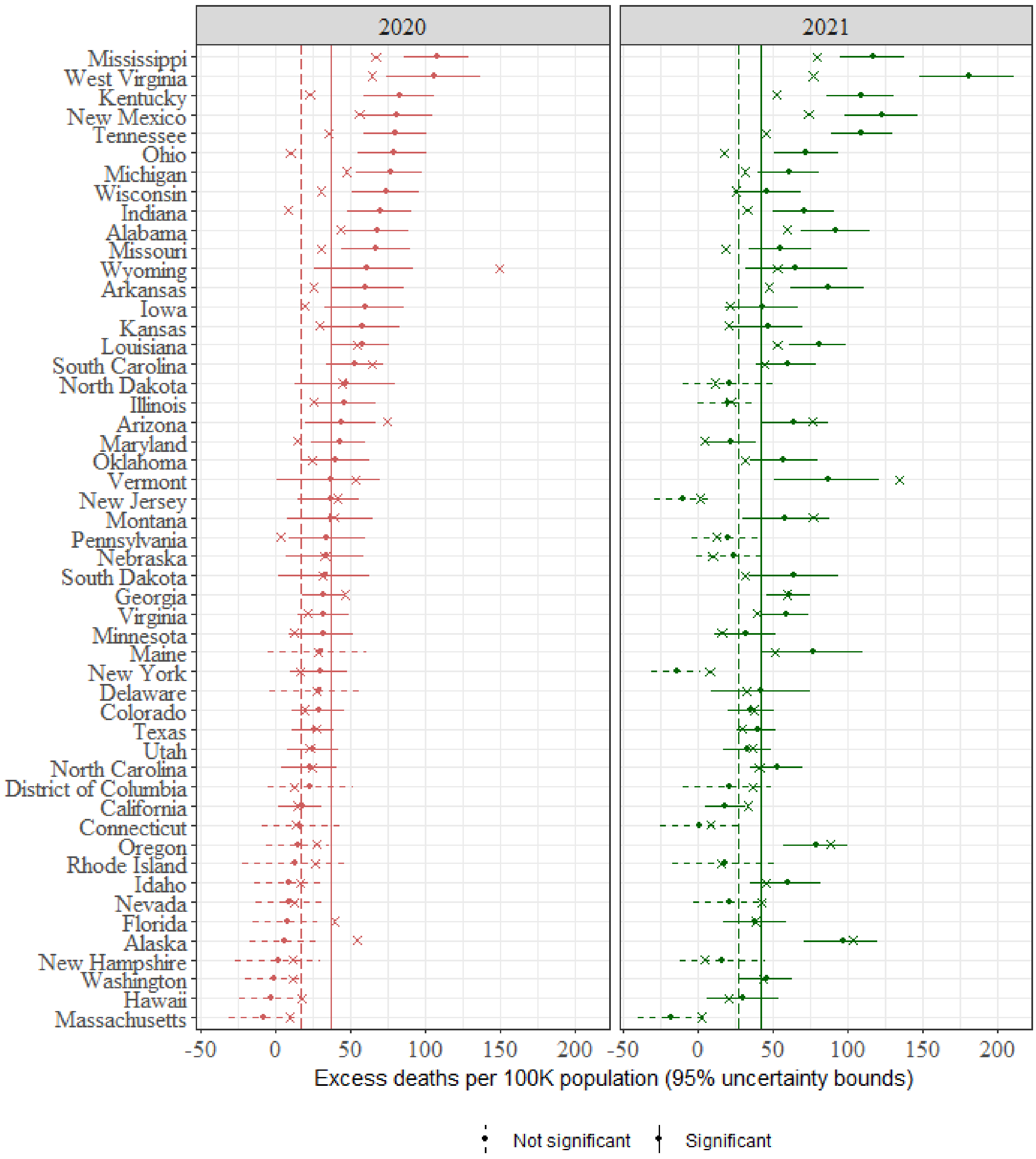


**Figure S2.** Comparison of excess mortality estimates for causes other than Covid-19, from NCHS and the current study. NCHS estimates are denoted by 'x' and our estimates are denoted by the point (median) and bars (95% uncertainty interval). Vertical lines denote the national excess estimate from this study (solid) and NCHS (dashed).

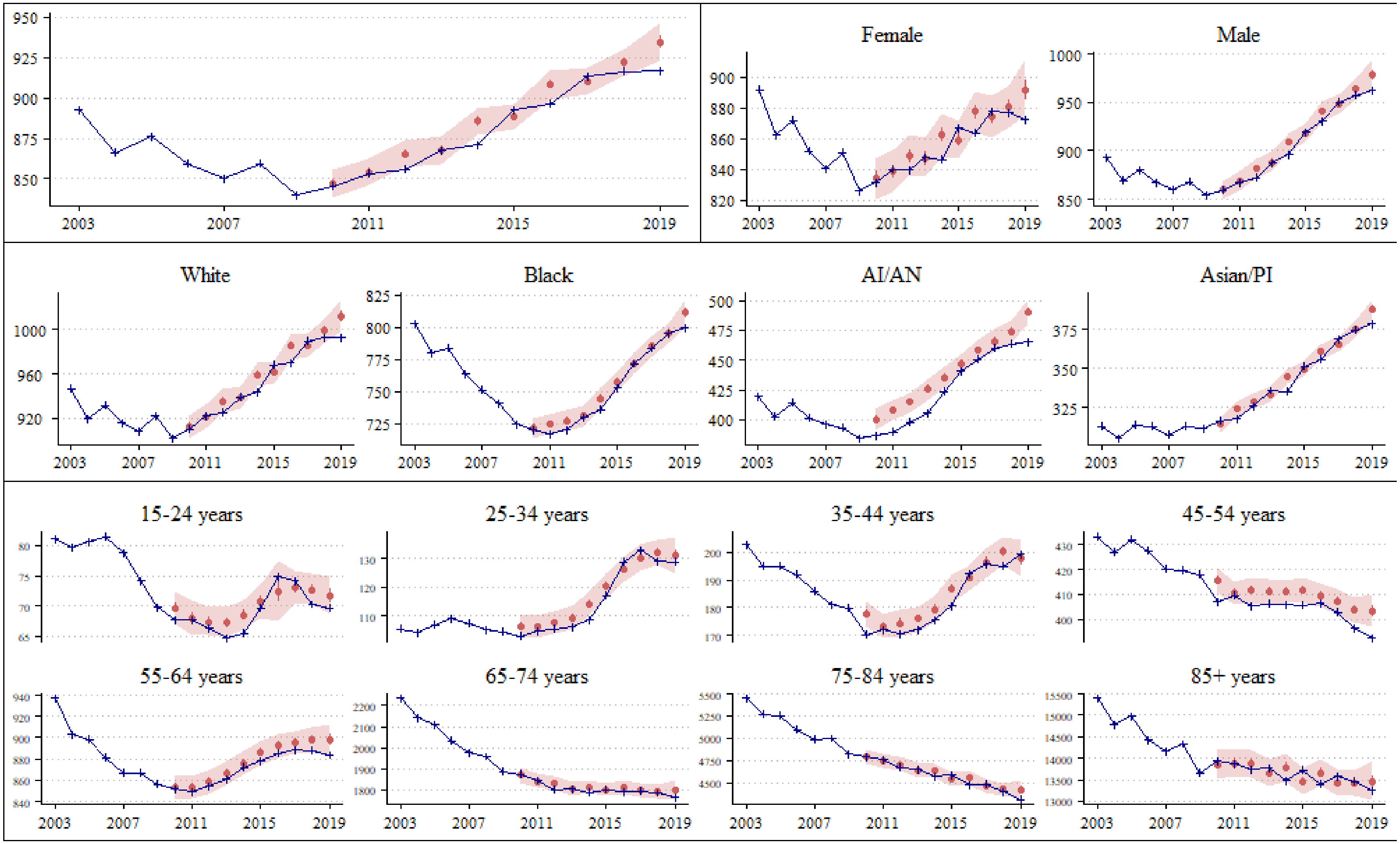


**Figure S3-a.** Cross validation of expected *all-cause* deaths for pre-Covid period 2010 – 2019, US national (top row, left), stratified by sex (top row, right), race (middle row) and age (bottom two rows). Observed deaths (in blue) and predicted median (red point), and 95% uncertainty interval (red shaded region) of expected deaths per 100,000 population.

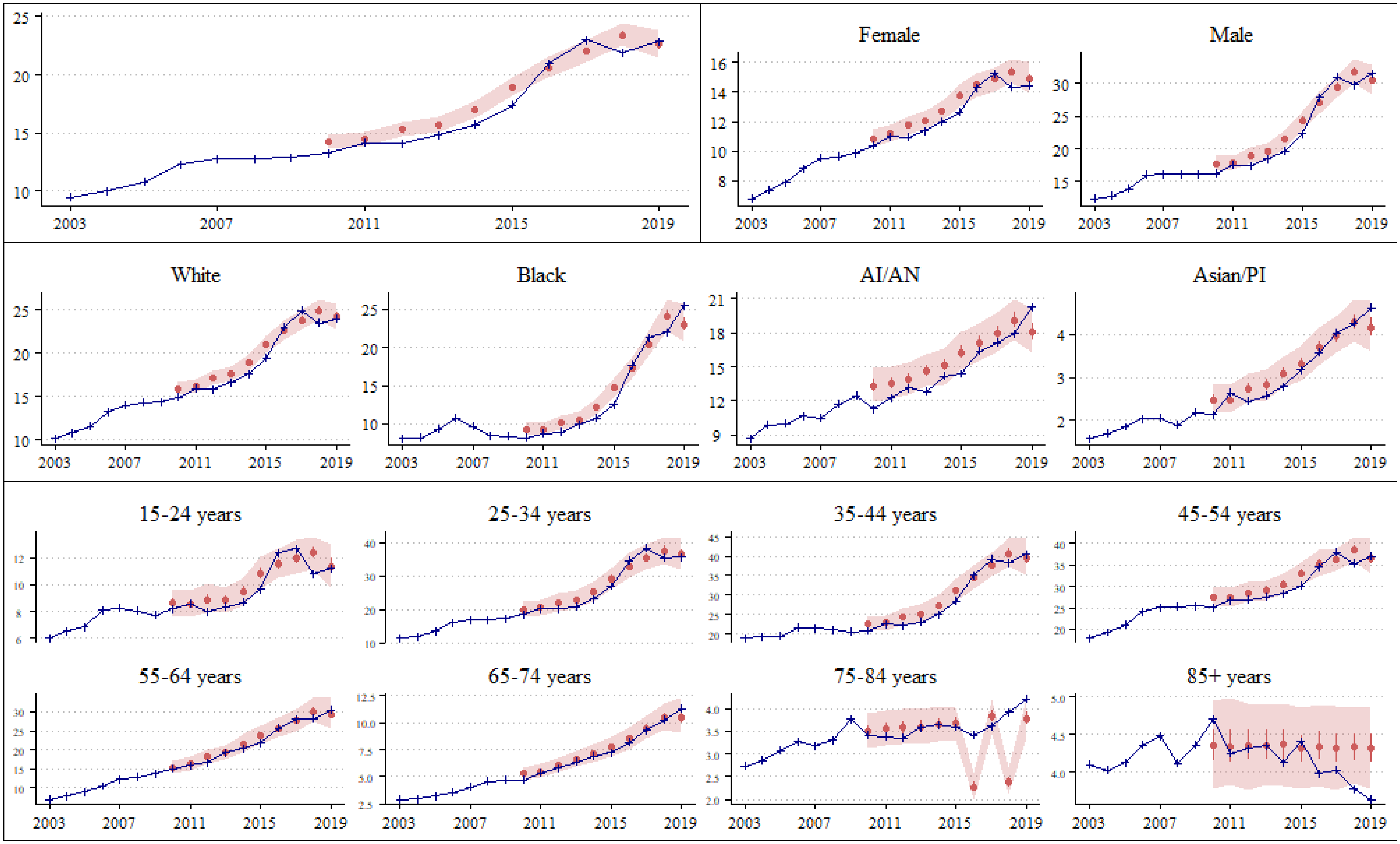


**Figure S3-b.** Cross validation of expected *poisoning* deaths for pre-Covid period 2010 – 2019, US national (top row, left), stratified by sex (top row, right), race (middle row) and age (bottom two rows). Observed deaths (in blue) and predicted median (red point), and 95% uncertainty interval (red shaded region) of expected deaths per 100,000 population.

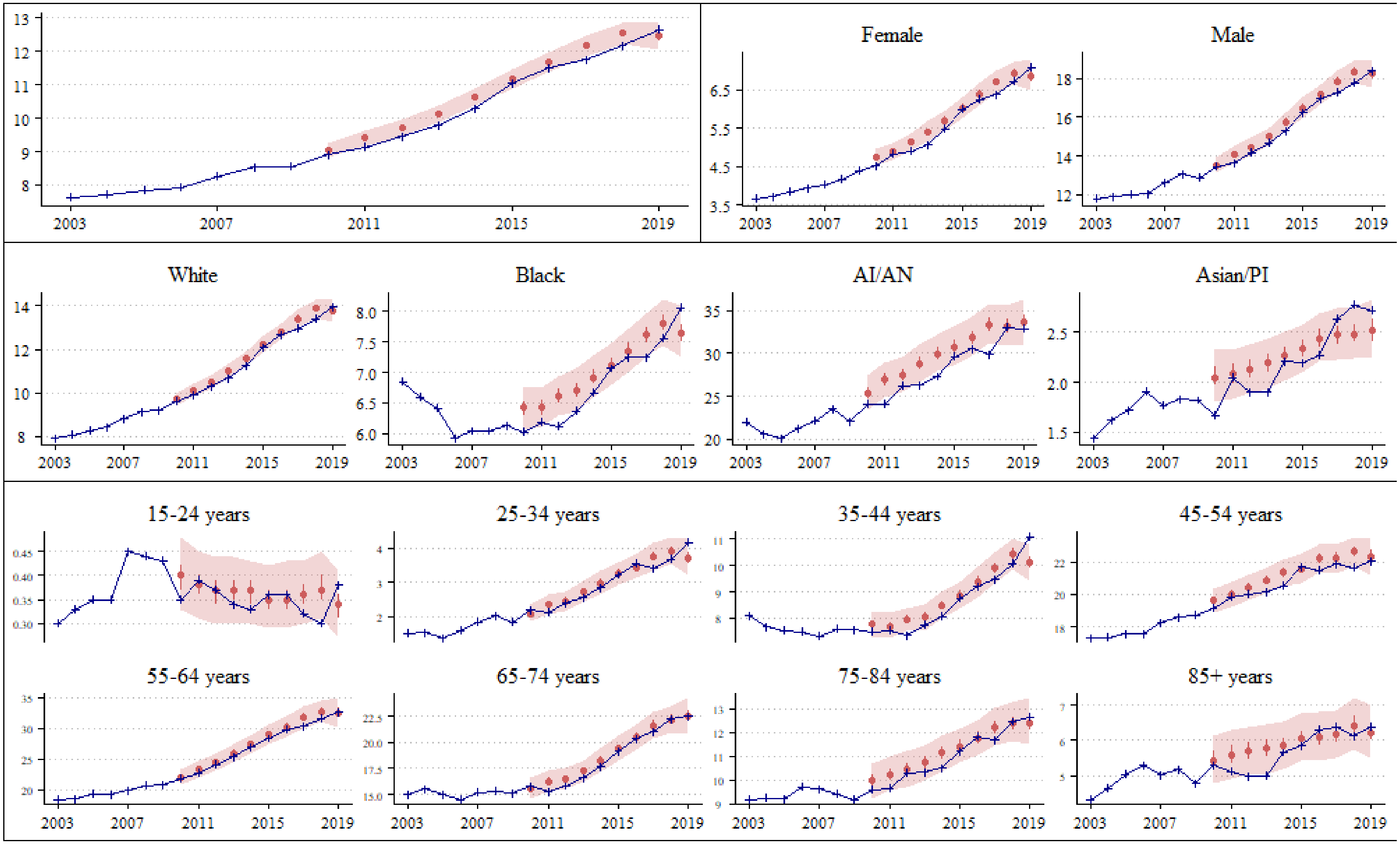


**Figure S3-c.** Cross validation for expected *alcohol-induced* deaths pre-Covid period 2010 – 2019, US national (top row, left), stratified by sex (top row, right), race (middle row) and age (bottom two rows). Observed deaths (in blue) and predicted median (red point), and 95% uncertainty interval (red shaded region) of expected deaths per 100,000 population.

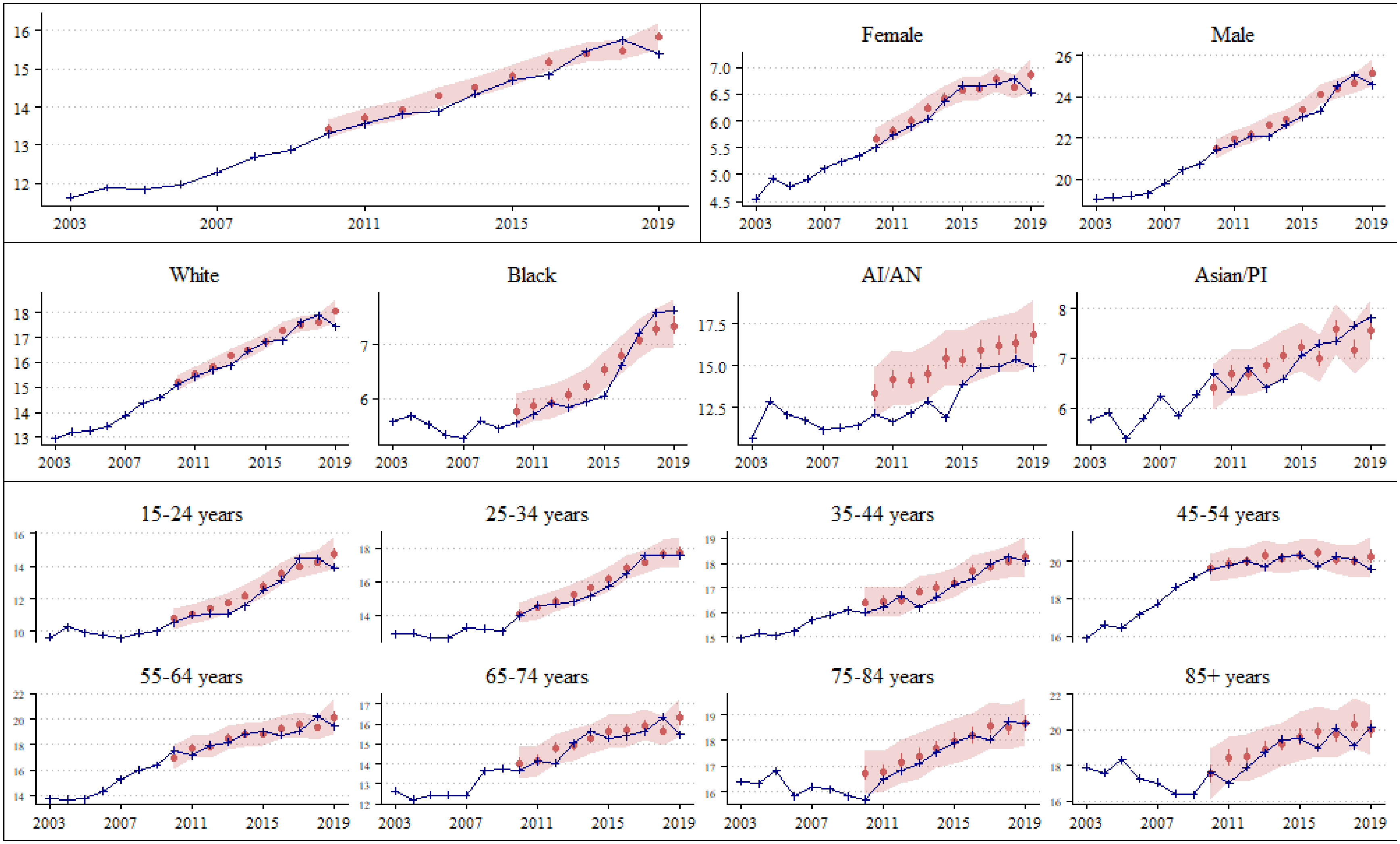


**Figure S3-d.** Cross validation for expected *suicide* deaths pre-Covid period 2010 – 2019, US national (top row, left), stratified by sex (top row, right), race (middle row) and age (bottom two rows). Observed deaths (in blue) and predicted median (red point), and 95% uncertainty interval (red shaded region) of expected deaths per 100,000 population.
